# Evaluation of a novel digital ostomy device on leakage incidents, quality of life, mental well-being, and patient self-care: an interventional, multicentre clinical trial

**DOI:** 10.1101/2024.06.10.24308691

**Authors:** Richard R.W. Brady, Diane Sheard, Mandie Alty, Martin Vestergaard, Esben Bo Boisen, Rachel Ainsworth, Helle Doré Hansen, Teresa Adeltoft Ajslev

## Abstract

**Background:** Most people with a stoma worry about leakage, and a quarter experience leakage of stomal effluent outside baseplate on a monthly basis. Leakage has additional physical and psychosocial consequences, for instance peristomal skin complications, feeling unable to cope and self-isolation.

**Method:** An interventional, single-arm, multi-centre, study was undertaken in United Kingdom, to evaluate a novel digital leakage notification system for ostomy care including a Support Service (=Test Product) for 12 weeks in patients with a recent stoma formation (≤9 months). Patients completed questionnaires at baseline and after 4, 6, 8, 10 and 12 weeks, evaluating leakage episodes, Ostomy Leak Impact (tool containing three domains) and patient self-management (by PAM-13). Additionally, mental well-being (by WHO-5) and quality of life (QoL) (by EQ-5D-5L) were assessed. Outcomes between baseline and final evaluation were compared by generalised linear- and linear mixed models.

**Results:** 92 patients (ITT population) were recruited with a mean age of 49.4-years (range 18-81 years). 80% had an ileostomy and 53% were female. After 12 weeks use of the Test Product, a significant decrease in mean episodes of leakage outside the baseplate (1.57 versus 0.93, *P*<0.046) was observed. Ostomy Leak Impact scores improved across all three domains (*P*<0.001), indicating less embarrassment, increased engagement in social activities, and increased control. Patient self-management also improved significantly (PAM-13 score: Δ6.6, *P*<0.001), as did the WHO-5 well-being index (Δ8.0, *P*<0.001). Lastly, EQ-5D-5L-profile-scores tended to improve (*P*=0.075).

**Conclusion:** A new digital leakage notification system demonstrated strong improvements to patients’ stoma self-care, mental well-being, and QoL.

**What does this paper add to the literature?:** Sensor technology embedded in supporting ostomy solutions can notify users about leakage seeping underneath the baseplate and thus secure a timely change of the baseplate before effluent may reach outside the baseplate soiling clothes or bedsheets. The technology showed potential in increasing quality of life for people with a stoma.

## Introduction

Around 200,000 people live with a stoma in the United Kingdom (UK) and approximately 13,500-21,000 undergo stoma surgery each year [1, 2]. Following stoma surgery patients must make significant adjustments to their normal life, including modification of lifestyle factors and behaviours, and need to accept an altered body image [3]. Many subsequently struggle with psychosocial and physical problems, e.g. depression, social isolation and peristomal skin complications (PSCs) [4, 5], leading to reductions in quality of life (QoL) [4, 6].

Early counselling and proactive teaching of stoma-management can enhance the psychological adjustment to stoma formation [7], however this may not always occur due to staff shortages and/or lack of time [8]. Even experienced users may struggle with self-care problems, including leakage of stomal effluent outside the ostomy solution (bag and baseplate), PSCs, frequent changes leading to high consumption of products, and long time needed for stoma care each day [5, 9]. Two out of three people with a stoma struggle with one or more self-care problems several years post-surgery [9].

Leakage of stomal effluent is the culprit of many of the problems experienced by people with a stoma [10–12]. Faeces located on the skin underneath the baseplate is an important risk-factor in the development of PSCs [11] and worry about leakage is associated with reduced emotional well-being and reduced engagement in social activities [10, 12]. Approximately two-thirds of respondents reported leakage outside baseplate at least once per year in a large global cohort [13].

People new with a stoma (<1 year since surgery) generally reported lower QoL compared with more experienced users, including reduced emotional wellbeing and social functioning, and impaired perception of body image [10, 14, 15]. One study also showed that a higher proportion of people new with stoma experienced weekly episodes of leakage outside baseplate compared with experienced users [10].

Since leakage of stomal effluent is a problem for people with recent stoma formation and continues to be a problem for many experienced users [10], it indicates that currently available products within stoma care do not sufficiently enable users to take proactive care to avoid leakage incidents progressing outside the baseplate and address the mental burden of worrying about leakage.

We recently reported results from an explorative clinical trial investigating a novel digital leakage notification system (Heylo™) as a stand-alone-solution for experienced users who struggled with leakage. The technical specifications of the system has previously been described, but in brief the system has been developed to help people with intestinal stomas gain better control of their stoma care by enabling users to know when effluent is seeping underneath the baseplate [16]. A *sensor layer* with two circular leakage sensor rings is placed between the baseplate and skin, and monitors for moisture as a sign of leakage. A *transmitter* attached to the *sensor layer* enables readout of the individual sensors and a *smartphone application* displays the state of the baseplate to the user. In the explorative study, Heylo™ reduced the number of leakage incidents progressing outside the baseplate, reduced worry about leakage and improved QoL [16].

Given the high levels of complications experienced in the early post-operative period in patients undergoing stoma formation, we in the present clinical trial tested Heylo™ delivered together with a Support Service in a population of patients who had undergone stoma formation within the last nine months in the UK. The aims of this study were to evaluate the effect of this technology on leakage, QoL and other outcomes, and to confirm and validate previous pilot study findings in this population.

## Methods

### Study design

The study was an interventional, single-arm, open-label, multicentre investigation enrolling patients to use Test Product (Heylo™ delivered together with a Support Service) for 12±2 weeks. A private clinical research organization (CRO) and nine National Health Service (NHS) hospital sites across UK recruited patients. Hospital sites recruited patients via chart/patient list reviews of those with recent surgery, independently of the Sponsor. The CRO recruited patients consecutively from a list provided to the CRO from Coloplast of patients registered within the Coloplast Charter database (Coloplast Charter offers support on products and routines and helps people with a stoma with product ordering and delivery) and who had stoma surgery within the past 9 months. All patients who completed this inclusion criteria were submitted to the CRO for further screening. Enrolment of patients at the CRO was independent from the Sponsor. The UK hospital sites were identified through open application and approached with assistance from the National Institute for Health and Care Research (NIHR) site identification processes.

Patients were invited for an information- and inclusion visit (V0) and signed consent forms before formally entering the study (Figure 1). Patients filled in questionnaires at baseline (V1) and after 4 (V2), 6, 8 (V3), 10 and 12 (V4) weeks use of Test Product. Study nurses conducted evaluations on contacts with healthcare professionals together with the patient at V1, V2, V3 and V4.

**Figure 1.**
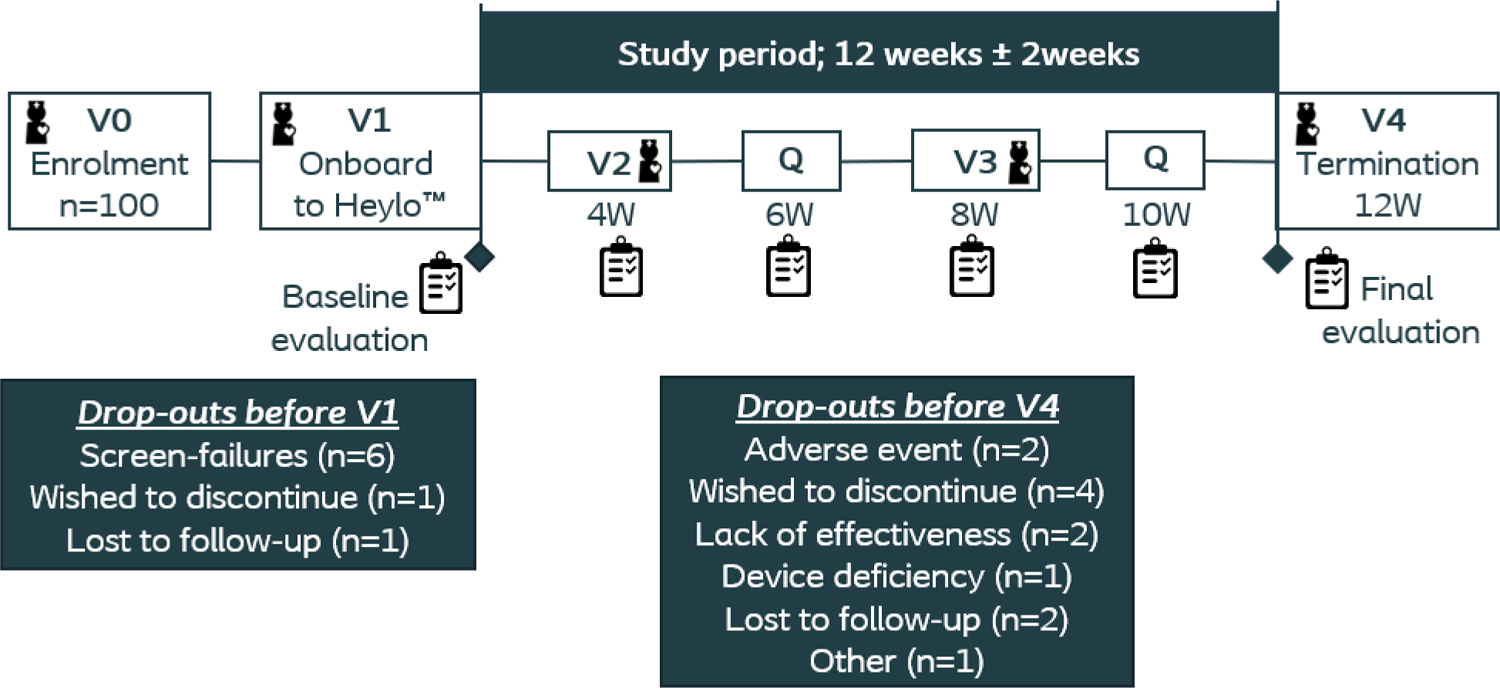
Overview of study design and drop-outs.

### Test Product (Heylo™ solution)

Patients were provided a Heylo™ starter kit (consisting of one transmitter, one charger and ten sensor layers), an additional transmitter, and enough sensor layers for users to change ostomy solutions to a similar routine as they would normally do. Patients were instructed to install the bespoke Heylo™ app on their personal smartphones. Heylo™ was delivered together with a remote Support Service (Coloplast Care Plus) consisting of three elements:

1. *Coloplast Charter* offers support on products and routines and helps people with a stoma with product ordering and delivery.
2. *Leakage Service*: Patients could call Coloplast Charter for support on leakage issues, and/or Coloplast Charter could reach out to patient based on triggers, if patient was struggling with leakage (observed from Heylo™ app leakage data in cloud).
3. Patient could call Coloplast Charter for *Technical Support (e.g. questions about Bluetooth connectivity)*.

The leakage notification system works both as a stand-alone solution and with the leakage support service included in this study. Availability of the leakage support service is currently country specific, nevertheless, a technical support service will be available in all countries.

### Selection of study participants

In respective sites, patients were identified by the research team from various sources, including, from operative and colorectal specialist multi-disciplinary team meeting lists and stoma care nursing databases. The sites recruited patients who were assessed to be able to follow study procedures for three months. Inclusion criteria identified those with an ileostomy or colostomy being >18 years and having liquid/mushy effluent (Bristol scale 5-7) [17]. Patients should have had their stoma for ≤9 months and have self-managed their stoma products for at least 14 days. Also, patients had to have a smartphone applicable to the bespoke Heylo™ app and be willing to sign up to Coloplast Charter (Dispensing Appliance Contractor) during the study, as other Dispensing Appliance Contractors currently cannot support the *Leakage Service* and *Technical Support*.

Patients could not be enrolled if they had stage 4 cancer and/or limited life expectancy. Patients with a complicated stoma at baseline (dehiscence/prolapse/hernia), with severe PSCs, and patients using topical steroid treatment in the peristomal area or receiving systemic steroid treatments were excluded. Patients with a pacemaker, known sensitivity to acrylate, and females being pregnant, or breastfeeding were excluded from participating in the study.

### Patient demographics and endpoints

Patient demographics were recorded at baseline.

At baseline and during test period patients filled in questionnaires evaluating various endpoints:

*Primary endpoint:*

- Self-reported number of events of stoma effluent leakage outside the baseplate within the past 2 weeks.

*Secondary endpoints:*

- Patient self-management using the Patient Activation Measure (PAM) instrument, 13-item version [18].
- Burden of leakage and QoL using the validated Ostomy Leak Impact (OLI) tool [19].
- Health-Related QoL (HRQoL) by the EQ-5D-5L [20, 21].

### Other assessment

- Assessment of psychological well-being by WHO-5, a five-item questionnaire [22].

Adverse events were recorded continuously throughout the study. Assessment of each adverse event and whether the adverse events were related to the Test Product was made and registered in the data management system by the study nurse and was afterwards independently assessed by the hospital site responsible Principal Investigator. All adverse events have been listed in Supplementary Table 1 and Supplementary Table 2.

### Statistics

Sample size calculation was based on a worst-case calculation, where the primary endpoint was evaluated as leakage outside the baseplate within the last 2 weeks (Yes/No) instead of using the exact number of times with leakage, since the distribution was unknown. If this proportion was reduced from 27% at baseline to 3% by end of study and using a 2-sided paired exact test in the binomial distribution (testing on a 5% level) we needed at least 45 patients to ensure a power of 85% to detect a significant difference. To allow for a dropout-rate of 25%, it was estimated that at least 60 patients should be enrolled. Statistical analyses were performed using SAS v. 9.4 (SAS Institute Inc., Cary, NC) after data entry and data management using a validated data management system (Smart-Trial version 2021.4.).

The primary endpoint was evaluated by paired comparison between data from 12 weeks (V4) (or last visit after at least 4 weeks use of Test Product if V4 data were missing) and baseline data (V1). A Poisson distribution was used for modelling data. The comparison was performed by a generalized linear mixed model with visit as a fixed effect, patient as a random effect and using a negative binomial distribution to allow for over-dispersion of the Poisson parameter.

The remaining endpoints were analysed like the primary endpoint except that they were assumed normally distributed and thereby were analysed by linear mixed models. Furthermore, the analyses included time since discharge as a covariate. The contribution of time since discharge to the baseline values was first inspected visually, and if evaluated applicable tested by linear regressions whether slopes were equal to zero.

### Ethical consideration

The study was carried out in accordance with the Declaration of Helsinki, ISO 14155:2011 and European Medical Device Regulation (2017/745) (MDR). The study was approved by the Ethical Committee of the *West Midlands - South Birmingham Research Ethics Committee* in UK before study initiation (IRAS Project-ID: 297458). The study was registered on ClinicalTrials.gov (*NCT05135754*). All patients were fully informed about the investigation, both verbally and in writing, and all gave written informed consent to participate in the study. Participation in the study was voluntary and patients could withdraw from the study at any time. The study was conducted from November 2021 to August 2022 in UK.

## Results

### Demographics of study participants

A total of 100 newly operated patients (≤9 months since stoma surgery) were enrolled in the study from ten sites across the UK (safety population), thus overrecruiting the intended number of patients. The hospital sites screened 187 patients of which 60 patients were enrolled (32.1%), and the CRO screened 325 patients of which 40 patients were enrolled (12.3%). Eight patients were omitted from the intention-to-treat (ITT) population and furthermore twelve patients did not complete the study as planned (Figure 1). Data from the ITT population (n=92) were included in the final statistical analyses.

Mean age of patients was 49.4 years (range 18-81; SD=14.7) and 53% were female. Eighty percent had an ileostomy and 20% had a colostomy. Reasons for stoma formation were cancer (34%), ulcerative colitis (22%) or Crohn’s disease (13%), or due to *other* causes (Table 1). On average, patients had their stoma surgery 140.9 days (range 21-275; SD=77.7) prior to enrolment.

**Table 1.** Demographics of intention-to-treat population.

| Parameter | Total (n=92) |
| --- | --- |
| **Age** (years): Mean ± SD (range) | 49.4 ± 14.7 (18; 81) |
| **Sex:** n (%) |  |
| Females | 49 (53.3%) |
| Males | 43 (46.7%) |
| **Days since stoma surgery** to V0: Mean ± SD (range) | 140.9 ± 77.7 (21; 275) |
| **Days since discharge from hospital** to V0: Mean ± SD (range) | 130.5 ± 76.1 (16; 256) |
| **Type of stoma:** n (%) |  |
| Ileostomy | 74 (80.4%) |
| Colostomy | 18 (19.6%) |
| **Reason for stoma creation\*:** n (%) |  |
| Ulcerative colitis | 20 (21.7%) |
| Cancer | 31 (33.7%) |
| Crohn's Disease | 12 (13.0%) |
| Other | 31 (33.7%) |
| **Ostomy solution brand\*\*:** n (%) |  |
| Coloplast | 61 (66.3%) |
| Another manufacturer | 33 (35.7%) |
| **Ostomy solution:** n (%) |  |
| 1-piece | 87 (94.6%) |
| 2-piece | 5 (5.4%) |
| **Baseplate type:** n (%) |  |
| Flat | 27 (29.3%) |
| Convex | 59 (64.2%) |
| Concave | 6 (6.5%) |

\* Two patients reported both a specific reason for stoma creation and pressed the option *other* as well.
\*\* Two patients reported using two different brands of ostomy solutions.

Two-third of the patients used Coloplast brand products as their regular ostomy solution and 95% of patients used 1-piece products. Moreover, two-third of patients used a convex product type (Table 1).

A total of n=108 calls with *Support Service* were recorded with the main points of discussion being leakage issues (58%) and questions related to Test Product (13%). Furthermore, n=29 *Technical Support* calls were conducted during the study period with the main points of discussion being issues with transmitter (40%) or Bluetooth connection (40%).

### Leakage outside baseplate

On average, patients experienced 1.57 (95%CI [1.19;2.08]) episodes of leakage outside baseplate in two weeks at baseline versus 0.93 (95%CI [0.56;1.54]) with Test Product, corresponding to a 41% reduction in episodes of leakage outside the baseplate (*P*=0.046) (Figure 2). Almost half of the patients (46%) did not report episodes of leakage outside the baseplate at baseline, this increased to 66% at the final evaluation.

**Figure 2.**
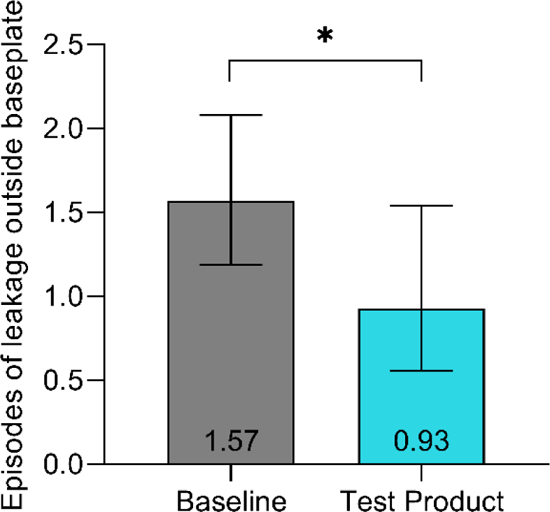
*Episodes of leakage outside baseplate.* Patients recalled episodes during the last 2 weeks. Data is presented as LS means and error bars represent the 95% confidence intervals. \**P*<0.05, \*\**P*<0.01, \*\*\**P*<0.001.

### Patient Self-management

PAM-scores improved on average Δ6.6 points (95%CI [3.45;9.78]) from 68.2 at baseline to 74.8 with Test Product (*P*<0.001) (Figure 3).

**Figure 3.**
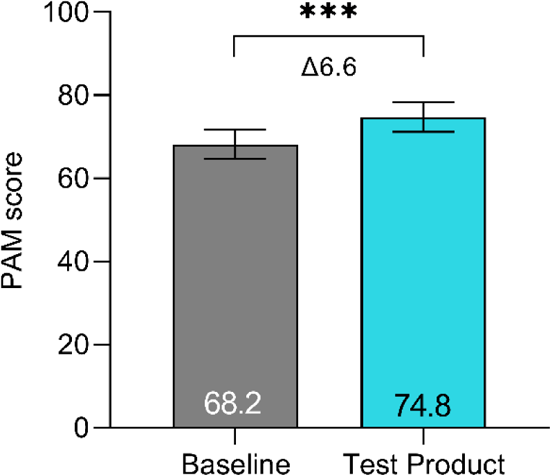
*Patient Activation Measure.* PAM scores at baseline and the final evaluation. PAM is scored on a scale ranging from 0 to 100. Individuals who score high on this instrument typically understand the importance of taking a proactive role in managing their health and have the skills and confidence to do so [18]. Data is presented as LS means and error bars represent the 95% confidence intervals. \**P*<0.05, \*\**P*<0.01, \*\*\**P*<0.001.

### Quality of life

Patients had significantly better scores in all three domains of the OLI tool when using the Test Product compared with baseline (Figure 4). The *Emotional impact* domain score increased with Δ20.0 points (95%CI [15.0;25.0], *P*<0.001), the *Usual and social activities* domain score increased Δ6.3 points (95%CI [2.7;9.9], *P*<0.001) and the *Coping and control* domain score increased Δ14.3 points (95%CI [8.3;20.4], *P*<0.001) from baseline to the final evaluation.

**Figure 4.**
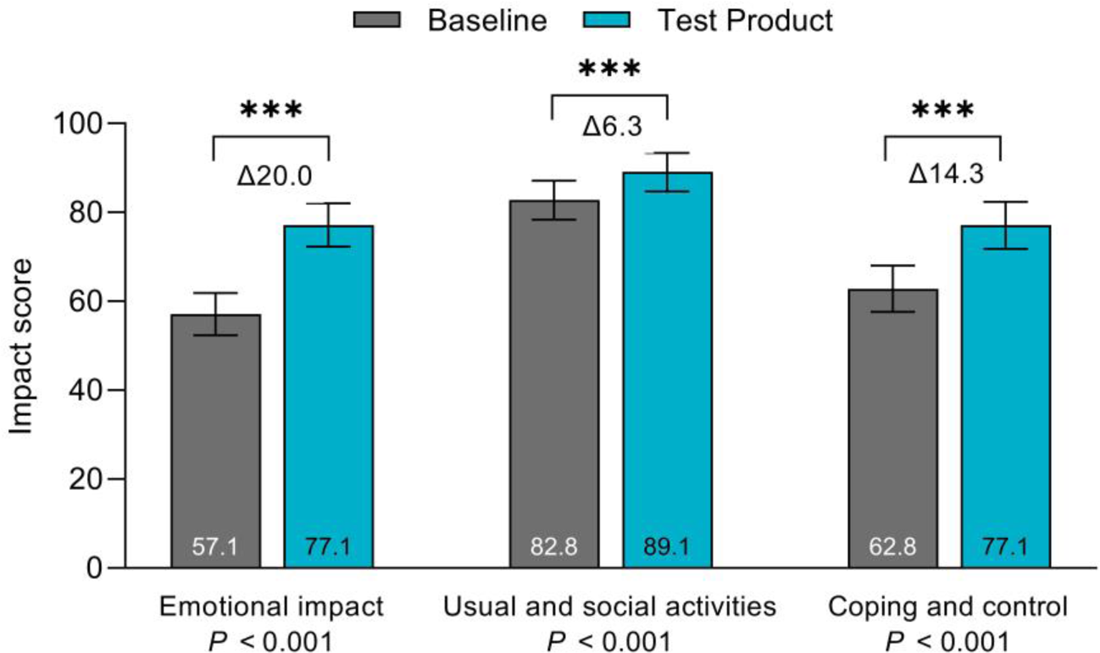
*Burden of leakage.* The OLI tool summarizes the burden of leakage in three domains: *Emotional impact*, *Usual and social activities* and *Coping and control*. Each domain sums into a total score ranging from 0 to 100. A higher score reflects lower impact [19]. Data is presented as LS means and error bars represent the 95% confidence intervals. \**P*<0.05, \*\**P*<0.01, \*\*\**P*<0.001.

Patients scored significantly higher on the generic WHO-5 well-being index, with the score increasing Δ8.0 points (95%CI [4.2;11.8], *P*<0.001) from 56.9 at baseline to 64.9 at the final evaluation (Table 2). Patients scored significantly higher on the EQ-5D-5L Visual Analogue Scale (VAS) with an improvement of Δ4.7 points (95%CI [1.6;7.8], *P*=0.004) from baseline to the final evaluation (Table 2). The EQ-5D-5L index score tended to increase (Δ0.034; 95%CI [-0.00;0.07], *P*=0.075) (Table 2).

**Table 2.** Evaluation of health-related QoL and mental well-being. Health-related QoL was assessed by the EQ-5D-5L questionnaire. Translation of health-states and index scores are based on the specific value set for UK. The second part of the questionnaire consists of a visual analogue scale (VAS) on which the patient rates perceived health from 0 (worst imaginable health) to 100 (best imaginable health) [20, 21]. Mental well-being was assessed using the WHO-5 questionnaire with the scale ranging from 0 (worst level of psychological well-being) to 100 (highest level of well-being) [22]. Data is presented as LS mean scores (95% Confidence Intervals).

| Parameter | Baseline | Final evaluation | Difference | P value |
| --- | --- | --- | --- | --- |
| **EQ-5D-5L** |  |  |  |  |
| Index score | 0.713 (0.663; 0.763) | 0.747 (0.696; 0.798) | 0.034 (-0.00; 0.07) | 0.075 |
| VAS score | 71.8 (68.1; 75.5) | 76.5 (72.7; 80.2) | 4.7 (1.6; 7.8) | 0.004 |
| **WHO-5** | 56.9 (51.9; 62.0) | 64.9 (59.8; 70.1) | 8.0 (4.2; 11.8) | <0.001 |

### Effect of time since discharge on outcome measures

It was assessed if time since hospital discharge had an impact on patients’ baseline outcome levels. As an example, for the *Emotional impact* domain of the OLI tool (Supplementary Figure 1), no significant change in baseline values were observed as a function of time since hospital discharge (*P*=0.933). Similar was observed for all other outcome measures, with none of the baseline outcome measures changing significantly as a result of time since hospital discharge (Table 3). These results were also reflected in the analyses of the endpoints, where the effect for the covariate (time since discharge) was not significant.

**Table 3.** Impact of time since discharge on baseline values of all outcome measures.

| Parameter | Slope (Score change / day) | P value |
| --- | --- | --- |
| Leakage outside baseplate | -0.00339 | 0.194 |
| PAM-13 score | -0.01275 | 0.522 |
| Emotional impact (OLI) | 0.00230 | 0.933 |
| Usual and social activities (OLI) | 0.01358 | 0.604 |
| Coping and control (OLI) | 0.03245 | 0.306 |
| EQ-5D-5L (Index) | 0.00028606 | 0.289 |
| EQ-5D-5L (VAS) | 0.00102 | 0.962 |
| WHO-5 | 0.00343 | 0.899 |

### Safety

In total n=88 adverse events were recorded in n=33 patients (33%), of these n=10 (n=5 patients; 5%) were serious. None (0%) of the serious adverse events, were independently assessed by the site based principal investigators to be related to the Test Product (Supplementary Table 1). A total of n=78 non-serious adverse events were recorded for n=29 patients (Supplementary Table 2). Twenty-one non-serious adverse events for n=18 patients were assessed by investigators to be ‘possibly’, ‘probably’, or ‘causally’ related to the Test Product. Most of the adverse events (n=20) were associated with skin and subcutaneous tissue disorders (primarily skin irritation) and one adverse event was recorded to be related to a gastrointestinal disorder (stoma bleeding). Intensity of two adverse events was considered moderate, and the remaining adverse events were considered mild. Two adverse events related to skin irritation caused two patients to discontinue the study. One patient discontinued the study due to a device deficiency (issue with transmitter), which could not have led to a serious adverse event. Consequently, no corrective actions were required to be taken.

## Discussion

Leakage of stomal effluent is a common problem for people with a stoma [13]. Currently available products within stoma care do not sufficiently enable users to take proactive care to avoid leakage progressing outside the baseplate and address the mental burden of worrying about leakage. Digital health solutions, i.e. wearable devices and connected healthcare solutions, are increasingly being adopted in healthcare systems across different areas of patient care [23].

In the present study, patients using a new digital leakage notification system experienced significantly fewer episodes of leakage outside the baseplate. By notifying patients about effluent seeping underneath the baseplate, this enabled them to inspect and change baseplate before effluent progressed outside the baseplate. The current study corroborates previous results from an exploratory investigation of the Test Product as a stand-alone solution for experienced users with leakage issues [16]. This indicates that the Test Product can help both patients newly discharged and experienced users in reducing number of leakages progressing outside the baseplate.

Previous studies have highlighted that the impact of leakage on participation in daily activities and the mental burden of worrying about leakage is correlated with the frequency of experiencing leakage episodes [10, 24]. In this study, patients scored significantly higher in all three domains of the OLI tool at the final evaluation compared with baseline (Figure 4). The magnitudes of the improvements were of clinical relevance with changes being similar to or higher than the minimally clinically important differences previously established (MCID-values based on average of three evaluation-methods: *Emotional impact* Δ7.6; *Usual and social activities* Δ6.6; *Coping and control* Δ7.2) [19], indicating that the Test Product provides a meaningful change for patients and overall means that patients felt less embarrassment, less frustration, better engagement in social activities, and felt better in control with their situation.

Moreover, patients scored significantly higher on the WHO-5 well-being index, with the score increasing Δ8.0 points from 56.9 at baseline to 64.9 at the final evaluation. The increase of Δ8.0 points on the WHO-5 scale was statistically significant, however may not necessarily be clinically relevant, since the MCID for this tool is described as a 10 percentage-point change [22, 25]. Nonetheless, after using Test Product for 12 weeks, the WHO-5 index well-being level reached the mean level of the general UK population, which in 2016 was 63.5 in those aged 35-50 [26], suggesting that the improvement observed had reached the baseline level for the UK population.

The EQ-5D-5L instrument was used to measure patients’ HRQoL, which can be used for economic evaluations and comparisons [27]. The baseline EQ-5D-5L index score was in the present study found to be 0.713 (UK specific), which is similar to scores for people with a stoma experiencing 1 to 4 leakage incidents per month reported in a time-trade-off study [28]. This is markedly lower than the score reported for the general population in England (0.885) [29]. In the present study, the EQ-5D-5L index score tended to increase from baseline to the final evaluation (Δ0.034, *P*=0.075). The baseline VAS score was 71.8 in this trial, thus, much lower than the mean self-rated VAS score of 82 found for the general population aged 45-54 years in UK [30]. After use of Test Product for 12 weeks the EQ-VAS score improved with Δ4.7 points to 76.5, though still in the lower end of the VAS level for the general population. Taken together, these data indicate that people with a stoma experience lower QoL and mental well-being compared with the general population and that use of the Test Product appeared to improve QoL and mental well-being in our study population.

The UK patient pathway for stoma care provides most support within the first year of stoma formation, where stoma care nurses try to empower patients to be able to self-manage their stoma care and subsequently the pathway recommends annual reviews concerning stoma management and product use [31]. An interesting, but nonetheless worrying observation was that for all endpoints, the baseline values across patients entering the study at different time points since hospital discharge were unchanged, indicating that patients entering the study nine months after stoma formation were not doing better than patients entering the study one month after stoma formation. This corroborates an earlier observation that people with recent stoma formation (<1 year) generally report higher burden of leakage compared with more experienced users [10] and may still be lacking basic support in stoma care provision, leading to increased pressure on the healthcare system. Clearly, new ways of supporting such patients may be required.

Supported self-management is part of the NHS Long Term Plan to empower people to better manage ongoing physical and mental health conditions themselves [32]. The PAM-13 tool was used to assess patient’s knowledge, skill, and confidence for managing their own health and healthcare [18]. The PAM-score improved on average Δ6.6 points when using Test Product, which is higher than the MCID of an at least 4-point difference [33]. This indicates that the Test Product provides a meaningful improvement in patients’ ability to manage their own health situation. High Patient Activation and self-management capability is associated with lower healthcare utilisation and less wasteful use of resources across primary and secondary care in UK [34, 35]. Indeed, a recent study highlighted that experiencing leakage incidents outside baseplate promoted behavioural changes leading to increased use of ostomy solutions, supporting products and interactions with health professionals, to mitigate the risk of future leakage events [24]. Supporting Patient Activation via digital solutions may potentially be a way to secure appropriate use of healthcare resources and ease the burden on the healthcare system. Future studies should identify the effect of this product and support service within specific populations, different types of stoma and for longer time-points. In addition, the role of more intensive follow-up of stoma pat*i*ents is an area of potential evaluation.

Study results should be interpreted considering limitations of the study design. The trial was a non-blinded, single-arm study, which might influence the subjective evaluations of the Test Product. Thus, improvements could be an effect of the Test Product, a study effect, due to natural improvements with time passing since surgery or a combination of all three factors. None of the endpoints were significantly changed as a function of time since hospital discharge. We therefore perceive that the improvements observed in the study are not a result of natural improvements over time, but due to an actual effect of the Test Product, with a potential influence by some study effect. Additionally, baseline values may be influenced by recall bias, since patients were not told to monitor leakage frequency until part of the study. The observed reduction in leakage episodes was lower than expected and likely influenced by several factors. Many patients did not experience leakage episodes outside their baseplate at baseline and a higher level than expected reported leakage episodes outside their baseplate when using the system. Since patients had a newly formed stoma (within 9 months of surgery), they may be more prone to episodes of sudden leakage incidents that, which reduce with experience and more stable behaviours. This may explain that leakage episodes did not reflect the 3% assumed in the sample size calculation. Moreover, for future investigations a more thorough understanding of the reported incidents should be explored, to report if these were or were not perceived as an ‘embarrassing situation’ for the patients (e.g. did the patient know that a leakage was on the way or did the patient experience fast progressing leakages that they could not react to in time?).

In conclusion, patients experienced significantly fewer leakage incidents outside the baseplate when using the Test Product and experienced significant improvements in QoL and mental well-being. Besides improving users’ QoL, patients also became more knowledgeable, pro-active, and engaged in managing their own health.

## Contributors

*TAA*, *EBB*, *RA* and *RRWB* conceptualized and designed the study. *RRWB* was the chief investigator of the study. *DS* and *MA* were involved in collection of data. *HDH* performed the statistical analyses. All authors had access to data in the study and all authors were involved in the interpretation of the data. *MV* wrote the first draft of the manuscript. All authors were involved in reviewing and editing of the manuscript. All authors gave final approval to publish the manuscript, and all agreed to be accountable for all aspects of the work.

## Data Availability

Deidentified data that underlie the results of this study, as well as study protocol, statistical analysis plan and informed consent form are available from the corresponding author upon reasonable request.

## Acknowledgement

The authors wish to express their sincere gratitude to all patients who participated in the study and to all health professionals involved in the study at the hospital sites (*The Newcastle Upon Tyne Hospitals NHS Foundation Trust*, *Leeds Teaching Hospitals NHS Trust*, *North Bristol NHS Trust*, *Chelsea & Westminster NHS Hospital*, *North West Anglia NHS Foundation Trust Peterborough City Hospital*, *James Paget University Hospitals NHS Foundation Trust*, *Royal Devon and Exeter NHS Foundation Trust*, *South Tees NHS Trust James Cook University Hospital*, and *Lancashire Teaching Hospitals NHS Trust Royal Preston Hospital*) and *Illingworth Research Group*.

## References

1. Kettle J. StoMap Programme Baseline Report. East of England NHS Collaborative Procurement Hub 2019

2. Aibibula M, Burry G, Gagen H, Osborne W, Lewis H, Bramwell C, et al. Gaining consensus: the challenges of living with a stoma and the impact of stoma leakage. Br J Nurs 2022; 31: S30–S9.

3. Sharpe L, Patel D, Clarke S. The relationship between body image disturbance and distress in colorectal cancer patients with and without stomas. J Psychosom Res 2011; 70: 395–402.

4. Richbourg L, Thorpe JM, Rapp CG. Difficulties experienced by the ostomate after hospital discharge. J Wound Ostomy Continence Nurs 2007; 34: 70–9.

5. Pearson R, Knight SR, Ng JC, Robertson I, McKenzie C, Macdonald AM. Stoma-related complications following ostomy surgery in 3 acute care hospitals: a cohort study. J Wound Ostomy Continence Nurs 2020; 47: 32–8.

6. Claessens I, Probert R, Tielemans C, Steen A, Nilsson C, Andersen BD, et al. The Ostomy Life Study: the everyday challenges faced by people living with a stoma in a snapshot. Gastrointest Nurs 2015; 13: 18–25.

7. O’Connor G. Teaching stoma-management skills: the importance of self-care. Br J Nurs 2005; 14: 320–4.

8. Di Gesaro A. Self-care and patient empowerment in stoma management. Gastrointest Nurs 2012; 10: 19–23.

9. Bulkley JE, McMullen CK, Grant M, Wendel C, Hornbrook MC, Krouse RS. Ongoing ostomy self-care challenges of long-term rectal cancer survivors. Support Care Cancer 2018; 26: 3933–9.

10. Jeppesen PB, Vestergaard M, Boisen EB, Ajslev TA. Impact of Stoma Leakage in Everyday Life: Data from the Ostomy Life Study 2019. Br J Nurs 2022; 31: S48–S58.

11. Voegeli D, Karlsmark T, Eddes EH, Hansen HD, Zeeberg R, Håkan-Bloch J, et al. Factors influencing the incidence of peristomal skin complications: evidence from a multinational survey on living with a stoma. Gastrointest Nurs 2020; 18: S31–S8.

12. Osborne W, White M, Aibibula M, Boisen EB, Ainsworth R, Vestergaard M. Prevalence of leakage and its negative impact on quality of life in people living with a stoma in the UK. Br J Nurs 2022; 31: S24–S38.

13. Martins L, Andersen BD, Colwell J, Down G, Forest-Lalande L, Novakova S, et al. Challenges faced by people with a stoma: peristomal body profile risk factors and leakage. Br J Nurs 2022; 31: 376–85.

14. Braumann C, Müller V, Knies M, Aufmesser B, Schwenk W, Koplin G. Quality of life and need for care in patients with an ostomy: a survey of 2647 patients of the Berlin OStomy-Study (BOSS). Langenbecks Arch Surg 2016; 401: 1191–201.

15. Davis JS, Svavarsdóttir MH, Pudło M, Arena R, Lee Y, Jensen MK. Factors impairing quality of life for people with an ostomy. Gastrointest Nurs 2011; 9: 14–8.

16. Brady RRW, Fellows J, Meisner S, Olsen JK, Vestergaard M, Ajslev TA. A pilot study of a digital ostomy leakage notification system: impact on worry and quality of life. Br J Nurs 2023; 32: S4–S12.

17. Lewis SJ, Heaton KW. Stool form scale as a useful guide to intestinal transit time. Scand J Gastroenterol 1997; 32: 920–4.

18. Hibbard JH, Mahoney ER, Stockard J, Tusler M. Development and testing of a short form of the patient activation measure. Health Serv Res 2005; 40: 1918–30.

19. Nafees B, Størling ZM, Hindsberger C, Lloyd A. The ostomy leak impact tool: development and validation of a new patient-reported tool to measure the burden of leakage in ostomy device users. Health Qual Life Outcomes 2018; 16

20. Feng Y-S, Kohlmann T, Janssen MF, Buchholz I. Psychometric properties of the EQ-5D-5L: a systematic review of the literature. Qual Life Res 2021; 30: 647–73.

21. Herdman M, Gudex C, Lloyd A, Janssen M, Kind P, Parkin D, et al. Development and preliminary testing of the new five-level version of EQ-5D (EQ-5D-5L). Qual Life Res 2011; 20: 1727–36.

22. Topp CW, Østergaard SD, Søndergaard S, Bech P. The WHO-5 Well-Being Index: a systematic review of the literature. Psychother Psychosom 2015; 84: 167–76.

23. Awad A, Trenfield SJ, Pollard TD, Ong JJ, Elbadawi M, McCoubrey LE, et al. Connected healthcare: Improving patient care using digital health technologies. Adv Drug Deliv Rev 2021; 178: 113958.

24. de Fries Jensen L, Rolls N, Russell-Roberts P, Vestergaard M, Jensen ML, Boisen EB. Leakage of stomal effluent outside the baseplate leads to rise in product usage and health professional interactions. Br J Nurs 2023; 32: 8–19.

25. Newnham EA, Hooke GR, Page AC. Monitoring treatment response and outcomes using the World Health Organization’s Wellbeing Index in psychiatric care. J Affect Disord 2010; 122: 133–8.

26. European Quality of Life Survey.

27. Wolowacz SE, Briggs A, Belozeroff V, Clarke P, Doward L, Goeree R, et al. Estimating health-state utility for economic models in clinical studies: an ISPOR good research practices task force report. Value Health 2016; 19: 704–19.

28. Rolls N, Yssing C, Bøgelund M, Håkan-Bloch J, de Fries Jensen L. Utilities associated with stoma-related complications: peristomal skin complications and leakages. J Med Econ 2022

29. Popping S, Kall M, Nichols BE, Stempher E, Versteegh L, van de Vijver DA, et al. Quality of life among people living with HIV in England and the Netherlands: a population-based study. Lancet Reg Health Eur 2021; 8: 100177.

30. Janssen M, Szende A, Cabases J, Ramos-Goñi JM, Vilagut G, König H-H. Population norms for the EQ-5D-3L: a cross-country analysis of population surveys for 20 countries. Eur J Health Econ 2019; 20: 205–16.

31. Davenport R. A proven pathway for stoma care: the value of stoma care services. Br J Nurs 2014; 23: 1174–80.

32. (Accessed: September 2023) NHS Long Term Plan.

33. Hibbard JH, Greene J, Tusler M. Improving the outcomes of disease management by tailoring care to the patient’s level of activation. Am J Manag Care 2009; 15: 353–60.

34. Barker I, Steventon A, Williamson R, Deeny SR. Self-management capability in patients with long-term conditions is associated with reduced healthcare utilisation across a whole health economy: cross-sectional analysis of electronic health records. BMJ Qual Saf 2018; 27: 989–99.

35. Bu F, Fancourt D. How is patient activation related to healthcare service utilisation? Evidence from electronic patient records in England. BMC Health Serv Res 2021; 21: 1–7.

