## Supplementary files for "Evaluation of a novel digital ostomy device on leakage incidents, quality of life, mental well-being, and patient self-care: an interventional, multicentre clinical trial"

**Supplementary Table 1**. Serious adverse events.

| | | |
|---|---|---|
| #1 | - Subarachnoid hemorrhage and encephalitis | Not related |
| #2 | - Low neutrophil count | Not related |
| #3 | - Bleeding stoma, potentially from food poisoning<br>- Dehydration, potentially from food poisoning<br>- Diarrhea, potentially from food poisoning<br>- Vomiting, potentially from food poisoning | Not related<br>Not related<br>Not related<br>Not related |
| #4 | - High output of black stoma contents | Not related |
| #5 | - Abdominal pain<br>- Intestinal blockage<br>- Intestinal infection | Not related<br>Not related<br>Not related |

**Supplementary Table 2**. Non-serious adverse events.

| | | |
|---|---|---|
| #1 | - Skin redness under arm of sensor layer, area red and itchy | Causal relationship |
| #2 | - Operation for injury on finger<br>- Skin redness 6 cm from stoma on the abdomen | Not related<br>Causal relationship |
| #3 | - Chest pain - diagnosed with Pulmonary Embolism<br>- Covid 19 Positive | Not related<br>Not related |
| #4 | - Abdominal pain and vomiting<br>- Generally unwell, lethargy, changes in urine, dehydration, output very liquid | Not related<br>Unlikely related |
| #5 | - Covid 19 Positive<br>- Headache<br>- Temperature | Not related<br>Not related<br>Not related |
| #6 | - Covid 19 Positive<br>- Parastomal Hernia | Not related<br>Unlikely related |
| #7 | - Soreness and bleeding around the stoma, on the hole. Also had this problem previously<br>- Red and itching skin beneath tail of Test Product | Unlikely related<br>Possibly related |
| #8 | - Small sore near stoma. It has been coming and going since stoma formation. | Unlikely related |
| #9 | - Red raw skin reported, painful and burning sensation, redness area reported as the size of stoma bag | Probably related |
| #10 | - Skin rash caused by the adhesive from the baseplate | Possibly related |
| #11 | - Skin irritation caused by the adhesive agent used on the baseplate | Causal relationship |
| #12 | - Feelings of urgency to have bowels open rectally<br>- Fatigue<br>- Itchy peristomal skin small area less than $10cm^2$<br>- Red itchy peristomal skin (whole circumference)<br>- Red peristomal skin, small area less than $10cm^2$ | Unlikely related<br>Not related<br>Probably related<br>Probably related<br>Probably related |
| #13 | - Red, itchy, dry under Test Product 'tail' | Probably related |
| #14 | - Dental extraction of tooth<br>- Headache<br>- Sore Throat<br>- Tooth pain<br>- Fatigue | Not related<br>Not related<br>Not related<br>Not related<br>Not related |
| #15 | - Itchy skin under transmitter | Possibly related |
| #16 | - Both cheeks on face itchy, red and dry<br>- Itchy skin underneath the 'tail' of Test Product | Not related<br>Causal relationship |
| #17 | - Skin redness is directly under the 'tail' of Test Product | Probably related |
| #18 | - Coryzal symptoms<br>- Diarrhea<br>- Fatigue<br>- Feels chilled/shivery<br>- Headache<br>- Headache<br>- Sore peristomal area under base plate | Not related<br>Unlikely related<br>Not related<br>Not related<br>Not related<br>Not related<br>Probably related |
| #19 | - Patient reports some discharge from the back-passage | Not related |
| #20 | - Diagnostic Sigmoidoscopy<br>- Patient reports to having some discharge coming through the back passage | Not related<br>Not related |

| | | |
|---|---|---|
| #21 | - Feeling of numbness in head when feeling panicky<br>- Headache<br>- Peristomal ulcer | Not related<br>Not related<br>Possibly related |
| #22 | - Small area of redness around perimeter of stoma | Probably related |
| #23 | - Arthritic pain in right wrist<br>- Headache<br>- Tension headache<br>- Redness around the edge of baseplate and stoma | Not related<br>Not related<br>Not related<br>Probably related |
| #24 | - Back Pain<br>- Hay fever<br>- Lower left back pain<br>- Redness at the underside of stoma crescent shaped | Not related<br>Not related<br>Not related<br>Probably related |
| #25 | - Back pain | Not related |
| #26 | - Cold sore<br>- Dehydration<br>- Exhaustion<br>- Flare up of eczema on right hand<br>- Increase of rectal discharge (blood and mucous)<br>- Inflammation of right wrist joint<br>- Injury to stoma<br>- Trauma injury to right foot<br>- Watery output from stoma<br>- Skin redness around the periphery of baseplate | Not related<br>Not related<br>Not related<br>Not related<br>Unlikely related<br>Not related<br>Not related<br>Not related<br>Not related<br>Possibly related |
| #27 | - Headache<br>- Headache | Not related<br>Not related |
| #28 | - Covid-19<br>- Feeling generally unwell<br>- Joint Pain in ankles and knees<br>- Joint Stiffness in ankles and knees<br>- Watery output from stoma<br>- Stoma bleeding<br>- Stoma increased redness | Not related<br>Not related<br>Not related<br>Not related<br>Unlikely related<br>Possibly related<br>Probably related |
| #29 | - Flexi sigmoidoscopy<br>- Pre-planned defaecating proctogram<br>- Stoma shrunk due to dehydration | Not related<br>Not related<br>Not related |

**Supplementary Figure 1**. Impact of time since hospital discharge on baseline values of the *Emotional impact* domain. Linear regression used to test if slope equal to zero.

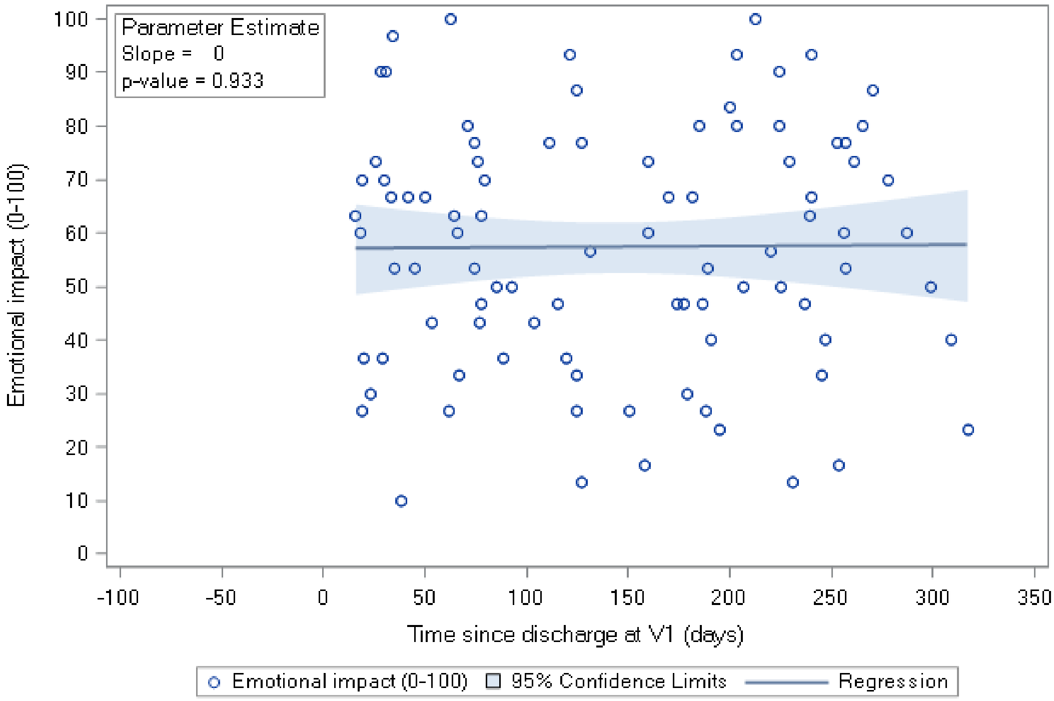
